# Global prevalence of antidepressant utilization in the community: A protocol for a systematic review

**DOI:** 10.1101/2022.02.17.22271152

**Authors:** Carlotta Lunghi, Michèle Dugas, Jacinthe Leclerc, Elisabetta Poluzzi, Cathy Martineau, Valérie Carnovale, Théo Stéfan, Patrick Blouin, Johanie Lépine, Laura Jalbert, Nataly R. Espinoza Suarez, Olha Svyntozelska, Marie-Pier Dery, Giraud Ekanmian, Daniele Maria Nogueira, Pelumi Samuel Akinola, Becky Skidmore, Annie LeBlanc

## Abstract

**Background:** Antidepressant drugs are the most frequently prescribed medication for mental disorders. They are also used off-label and for non-psychiatric indications. Prescriptions of antidepressants have increased in the last decades, but no systematic review exists on the extent of their use in the community.

**Methods and analysis:** We will conduct a systematic review to estimate the prevalence of antidepressant use in the community. We will search for studies published from 2010 in the Embase and MEDLINE databases. Study selection (by title/abstract and full-text screening) and data extraction for included studies will be independently conducted by pairs of reviewers. We will then synthesize the data on the prevalence of antidepressant use in individuals living in the community. If possible, we will perform a meta-analysis to generate prevalence-pooled estimates. If the data allows it, we will conduct subgroup analyses by antidepressant class, age, sex, country or other sociodemographics. We will evaluate the risk of bias for each included study through a quality assessment using the Joanna Briggs Institute Critical Appraisal tool: Checklist for Studies Reporting Prevalence Data. DistillerSR software will be used for the management of this review.

**Ethics and dissemination:** Ethical approval is not required for this review as it will not involve human or animal subjects. The findings of our systematic review will be disseminated through publications in peer-reviewed journals, the Qualaxia Network (https://qualaxia.org), presentations to international conferences on mental health and pharmacoepidemiology, as well as general public events.

**PROSPERO registration details:** CRD42021247423

## INTRODUCTION

Of the roughly 800 million people worldwide with a mental disorder, depression and anxiety are the most frequent, and both have a significant burden of disability (1). Antidepressants are first-line medications to treat current mental disorders, such as depression and anxiety (2-4), and these indications are those driving the number of prescriptions (5). Nevertheless, these medications are also prescribed for other in-label and off-label indications such as insomnia, pain, fibromyalgia, eating disorders, smoking cessation, migraine, and attention-deficit/hyperactivity disorders (5-10).

In the last two decades, various epidemiological studies have shown an increased prevalence of antidepressant prescriptions in industrialized countries (11-17). This could be due to an increased prevalence of current mental disorders (18, 19), which may also be due to primary care physicians’ improved ability to recognize these disorders and promptly begin pharmacological treatment. Conversely, other studies suggest a relatively stable prevalence of mental disorders or under-recognition and undertreatment (20, 21). Other facilitating factors possibly contributing to the rise in antidepressant prescriptions and use are the availability of new medications with a better risk-benefit profile (e.g., selective serotonin receptor inhibitors - SSRIs) (22), the introduction of generics on the market (23), experience or fear of withdrawal symptoms (24), other socioeconomic and cultural factors (e.g., stigma mental health well-being campaigns)(25, 26), or increased duration of treatment (27, 28).

A Canadian study on the surveillance of antidepressant drug prescription patterns showed an increased prevalence between 2006 and 2012, from 9% to 13% (29). Nevertheless, the incidence rate remained approximately stable in the same period (29). Similar data on the incidence and prevalence of antidepressant utilization were also reported by other studies in different countries (11, 13, 16, 27, 28). Thus, these results may indicate that the rise in prevalence could be due, at least partially, to an increased mean treatment duration rather than a higher number of patients being prescribed antidepressants. Indeed, a Finnish study estimated that, among antidepressant users in 2000-2001, 43% were long-term users, 32% intermittent, and only 26% short-term users. Moreover, only three-quarters of them had a psychiatric condition for which an antidepressant would have been appropriate (30). A more recent study conducted in Italy showed that almost 30% of patients who started an antidepressant drug treatment in 2013 were still on medication three years later (31). Among them, 10% used more than 180 defined daily doses (DDDs) per year (31). In addition to these significant changes in prescriptions and use over time, the prevalence in antidepressant drug use also varies according to age (12, 14), sex (12), country (14, 25, 32, 33) and antidepressant agent or class (17, 32, 34).

Despite the extensive utilization of antidepressant drugs worldwide, the increased use of the last decades, and the differences according to relevant sociodemographic factors, no systematic review exists on the prevalence of antidepressant use in the community. To our knowledge, the only systematic reviews on the use of antidepressants focused on specific populations, such as pregnant women (35) or people with particular diseases, such as cancer (36) or acute coronary syndrome (37). Estimating the prevalence of antidepressant utilization in the general population is essential to inform researchers, clinicians and decision-makers on prescription patterns over time and according to age groups and sex to guide new research, clinical decisions and allocation of health resources. Surveillance of antidepressant use may thus highlight potentially inappropriate prescriptions, such as their use in mild depression (38). Therefore, this systematic review aims to estimate the prevalence of antidepressant use among children and adolescents, adults and older adults living in the community.

## METHODS AND ANALYSIS

We will conduct a systematic review following the JBI Manual for Evidence Synthesis (39) for its conduct and the PRISMA (40) and MOOSE (41) guidelines for its reporting. The current protocol has been published in the International Prospective Register of Systematic Reviews database (PROSPERO no. CRD42021247423).

We have engaged with a panel of knowledge users (patients, caregivers, clinicians) and researchers in establishing our review question and literature search strategy. We will continue to engage them through the review process (e.g., data extraction, results interpretation, and findings dissemination).

### Participants

We will include studies on participants living in the community and exposed to antidepressants, independently of age, sex, ethnicity, religion or geographical area. We will exclude all the studies focusing on inpatient populations only (e.g., hospitalized patients, nursing homes) and those focusing on patients with a specific disease (e.g., depression or cancer), condition (e.g., pregnant women) or from a particular social group (e.g., health care workers, veterans).

### Exposure

We will include studies reporting on antidepressant use independently of class. Thus all will be included: SSRIs, Serotonin and Norepinephrine Reuptake Inhibitors (SNRIs), Monoamine Oxidase Inhibitors (MAOIs), Tricyclic Antidepressants (TCAs) and atypical antidepressants.

### Outcomes

The primary outcome will be the prevalence of antidepressant use.

### Study design

We will include studies with a descriptive observational design reporting the prevalence of antidepressant use (e.g., cohort studies, cross-sectional studies). Experimental, quasi-experimental, case-series and case-reports studies will be excluded. Case-control studies will be included only if the control group is representative of the general population. We will exclude reviews, commentaries, editorials, letters to the editor, lectures, theses, conference abstracts and grey literature.

### Language

No language restriction will be applied.

### Search strategy

Search strategies were developed by an experienced medical information specialist (BS) in collaboration with the research team and knowledge users during the protocol phase to ensure feasibility. The MEDLINE strategy was peer-reviewed by a second information specialist following the PRESS checklist. For the search, we used a combination of controlled vocabulary (e.g., “Antidepressive Agents”, “Incidence”, “Drug Utilization”) and keywords (e.g., “antidepressants”, “SSRI”, “prevalence”). We will search Embase and MEDLINE (including Epub Ahead of Print and In-Process & Other Non-Indexed Citations) and adjust vocabulary and syntax across databases. We will then download results and eliminate duplicates using EndNote version 9.3.3. (Clarivate). We decided to limit our results to the publication years from 2010 to the present. The rationale for this choice was to provide the most up-to-date evidence regarding antidepressant use. Additionally, with antidepressant use increasing in recent years, this strategy minimizes the risks of underestimating its prevalence.

### Study selection and data extraction

We have developed standardized forms to select eligible studies through title and abstract screening and full-text examination, and we will conduct pilot testing of each form across all reviewers. Pairs of reviewers will independently undertake title, abstract, and full-text screening and data extraction. Discrepancies between reviewers will be resolved by discussion or arbitration of a third senior reviewer. Extracted data will include (1) Study identification (e.g., title, journal, year of publication); (2) Study characteristics (e.g., country, study design, source of data); (3) Population characteristics (e.g., age, gender, ethnicity,); and (4) Outcomes (e.g., prevalence, indication/diagnostic, drug prescribed). We will use the DistillerSR software for the management of this review (DistillerSR. Version 2.35. Evidence Partners; 2021. Accessed April 2021-February 2022. https://www.evidencepartners.com).

### Quality assessment

Pairs of reviewers will independently assess the methodological quality of the included articles using the Joanna Briggs Institute Critical Appraisal tool: *Checklist for Studies Reporting Prevalence Data*. All the discrepancies between reviewers will be resolved by discussion or arbitration of a third senior reviewer.

### Data synthesis and analysis

We will synthesize the data on the prevalence of antidepressant drug utilization. Where possible, we will conduct subgroup analyses according to different relevant variables reported in the selected studies. Particular attention will be placed on age groups (children and adolescents; young adults; adults; and older adults) and sex differences since antidepressant use (and diseases for which antidepressants are prescribed) varies significantly according to these characteristics (12, 14, 42). If relevant, other subgroup analyses will be explored, such as antidepressant class, country, socioeconomic status, or ethnicity. We will undertake a meta-analysis to generate estimates of antidepressant use prevalence across included studies if the data allows it. Subgroup and sensitivity analyses will be performed when possible and appropriate. If a meta-analytic approach is possible, we will use the I^2^ statistic to evaluate heterogeneity. In case of heterogeneity (i.e., I^2^>50%) across studies, we will use random-effects models. An experienced biostatistician of the group will conduct the meta-analyses.

## EXPECTED LIMITATIONS

This systematic review protocol may have a few limitations. First, despite the extensive databases search, we did not include grey literature in the search strategy. Moreover, we may not be able to perform a meta-analysis, depending on the available data. In fact, a pooled estimation of the prevalence of antidepressant drug use will be valid only if the heterogeneity among studies is not too large. Differences in populations, data sources, study designs and antidepressants studied may thus preclude a meta-analysis.

## ANTICIPATED RESULTS

Drug utilization studies are essential to highlight prescription practices and uses of drugs in a real-world context. Nevertheless, systematic reviews of drug utilization studies are missing, except for a few specific populations or diseases. This review will be the first to synthesize information on the global extent of antidepressant use in the community. We will provide evidence on the epidemiology of antidepressant drug utilization over the last decade and differences between age groups and sexes. Variability across countries, databases and health systems will be reported too. We will analyze results on antidepressant use in light of current clinical guidelines for antidepressant primary indications (e.g., depression and anxiety). Clinical practice guidelines are essential for clinicians to decide when to start an antidepressant, which drug to prescribe, and how long to continue the treatment, all depending on patient characteristics. Thus, this systematic review will contribute to the knowledge on antidepressant use among different patients subgroups. It may allow for highlighting their possible inappropriate use in terms of drug type, duration of treatment, indication or patient characteristics (i.e., frailty elders), according to the availability of the information. The evidence will guide clinicians when prescribing these drugs, improving the quality of care offered to people with mental disorders. The results may also guide governments when designing public health policies in mental health, especially to promote, prevent or treat common mental disorders, such as depression and anxiety.

## Data Availability

All data produced in the present work are contained in the manuscript

## ETHICS AND DISSEMINATION

This systematic review does not require ethical approval since it will not involve human or animal subjects. Preliminary results of this systematic review will be presented to the patient partner and knowledge users (Qualaxia Network representatives) to involve them in interpreting and understanding the potential implications of the results and getting their feedback. We will produce a dissemination report for the knowledge users and share the results on social media platforms and through webinars for researchers and healthcare professionals of Quebec. A special issue on the Qualaxia Network Website will cover the results of this systematic review. In addition, a short and standardized policy brief will be shared through the SPOR Evidence Alliance Website. We will further disseminate results through presentations at scientific conferences, research webinars and manuscripts submitted to scientific, peer-reviewed journals for publication.

## AUTHORS’ CONTRIBUTIONS

CL, EP and JaL initially conceived the study. AL, BS, CL, JaL, JoL, and MD substantially contributed to the design of the study methods. CL, JoL and MD prepared the PROSPERO submission. AL, BS, CL, JaL, JoL, and MD elaborated the search strategy, and BS will perform the databases searches. AL, CL, CM, DMN, GE, JoL, LJ, MD, MPD, NE, OS, PB, PSA, TS and VC will perform the screening selection by title and abstract. AL, CL, CM, GE, JoL, MD, NE, OS, PB, TS, and VC will perform the screening selection by full-text examination. CL and CM produced the first draft of this manuscript. AL, BS, DMN, EP, GE, JaL, JoL, LJ, MD, MPD, NE, OS, PB, PSA, TS, and VC critically commented on the first draft and substantially contributed to the final version. All the authors approved the final version of this protocol.

## ACKNOWLEDGMENTS

We thank Kaitryn Campbell, MLIS, MSc (St. Joseph’s Healthcare Hamilton/McMaster University), for the MEDLINE search strategy peer review.

